# Avoidant restrictive food intake disorder (ARFID) in Swedish preschool children: a screening study

**DOI:** 10.1101/2024.09.26.24314270

**Authors:** Lisa Dinkler, Katarzyna Brimo, Helena Holmäng, Kahoko Yasumitsu-Lovell, Ralf Kuja-Halkola, Anne-Katrin Kantzer, Zerina Omanovic, Narufumi Suganuma, Masamitsu Eitoku, Mikiya Fujieda, Elisabeth Fernell, Per Möllborg, Rachel Bryant-Waugh, Christopher Gillberg, Maria Råstam

## Abstract

**Background:** Despite its common early onset, little is known about the prevalence and clinical presentation of avoidant restrictive food intake disorder (ARFID) in very young children, hindering early identification and intervention. Differentiating ARFID from normative selective eating is particularly challenging, yet validated parent-reported screening tools are lacking. This study aimed to estimate the point prevalence and describe the clinical characteristics of ARFID in preschoolers. It also evaluated the psychometric properties of the parent-reported ARFID-Brief Screener by assessing its agreement with a diagnostic interview for ARFID.

**Methods:** Parents of 645 children (50.5% male, mean age 3.2 years) completed the ARFID-Brief Screener and a neurodevelopmental screener during 2.5- and 4-year routine check-ups at 21 child health centers in West Sweden. Parents of all screen-positive and of randomly selected screen-negative children were invited to a follow-up diagnostic interview via phone. Additional clinical data were extracted from health records.

**Results:** Of the 42 children (6.5%) who screened positive for ARFID, 29 were followed up via diagnostic interview, and 21 received an ARFID diagnosis, yielding a positive predictive value of 72%. Negative predictive value, sensitivity, specificity, and overall accuracy of the ARFID-Brief Screener were 94%, 91%, 79%, and 84%, respectively. The estimated point prevalence of ARFID was 5.9%. All diagnosed children exhibited both sensory-based avoidance and low interest in eating. Only 13.5% met ARFID criteria based on weight- or nutrition-related impairment (DSM-5 Criteria A1-A3). Two fifths (39.1%) of children with ARFID exhibited early language delays compared to 13.5% of children without ARFID. More extensive neurodevelopmental problems were associated with greater ARFID severity and with higher scores on the sensory and concern profiles.

**Conclusions:** ARFID is not uncommon among preschoolers, though prevalence may be slightly overestimated in this study. It is primarily characterized by sensory-based avoidance and low interest in eating, and by psychosocial impairment instead of physical health consequences, underscoring the need to assess impact beyond weight, growth, and nutrition. Early neurodevelopmental difficulties are overrepresented, highlighting their relevance for early detection and intervention. The ARFID–Brief Screener demonstrated promising psychometric properties and may be a valuable tool for routine screening, though follow-up assessments remain necessary to confirm a diagnoses.

**Plain English summary:** Avoidant restrictive food intake disorder (ARFID) often starts early in life, but little is known about how common it is in very young children. This study looked at how many preschool-aged children have ARFID and what their eating difficulties look like. We also tested how well a short parent questionnaire, the ARFID-Brief Screener, can help identify children at risk.

Parents of 645 Swedish children filled out the screener during routine health check-ups at age 2.5 or 5. Children who screened positive and a randomly selected group of screen-negative children were invited to a follow-up interview. Based on these interviews, we estimated that about 5.9% of preschoolers may have ARFID. All children with ARFID avoided food due to sensory sensitivities or had little interest in eating. Few had weight or nutrition problems, but many experienced difficulties in daily life, such as stressful mealtimes.

Children with ARFID were more likely to have early neurodevelopmental challenges such as delayed language development, suggesting that these symptoms may aid in identifying children at risk for ARFID. The ARFID-Brief Screener performed well in detecting at-risk children, but follow-up assessments are still needed to confirm a diagnosis.

## 1. Introduction

Avoidant restrictive food intake disorder (ARFID) is a feeding and eating disorder characterized by severe limitations in food intake, leading to significant nutritional, medical, and psychosocial impairments (American Psychiatric Association, 2013; World Health Organization, 2022). Unlike other eating disorders, food restriction in ARFID is not primarily driven by preoccupation with body weight or shape but rather by *Low interest* in food, *Concern* about aversive consequences (e.g., choking, vomiting), and/or *Sensory* aversions (e.g., smell, taste, texture). Additionally, ARFID typically has an earlier onset, often in early or middle childhood, particularly in the Low interest and Sensory profiles (American Psychiatric Association, 2013).

Despite the common early onset, little is known about the prevalence and clinical presentation of ARFID in very young children, hindering effective healthcare planning and resource allocation for this group. Since its inclusion in DSM-5, several screening studies have assessed ARFID prevalence in the general population across various countries, genders, and age groups; however, few studies have focused on children under six (Sanchez-Cerezo et al., 2023). Identifying ARFID in preschoolers is particularly complex because selective (“picky”) eating is common in children under six and is considered part of normative development, typically resolving without intervention by age six or seven (Bourne et al., 2023; Breiner et al., 2024; Cardona Cano et al., 2015). This makes distinguishing ARFID from typical selective eating challenging, increasing the risk of misdiagnosis. Normative selective eating might be prematurely labeled as ARFID, potentially overburdening healthcare resources. Conversely, parents of children with severe restrictive eating may be reassured that this issue is temporary, missing the opportunity for early identification and treatment of ARFID, which could lead to a chronic, potentially lifelong disorder (Breiner et al., 2024).

To differentiate ARFID from normative selective eating, it is important to identify clinically significant impairment by assessing any detrimental nutritional, medical, or psychosocial impacts of the eating behavior as outlined by DSM-5 and ICD-11. While evaluating the *physical* consequenes of ARFID (e.g., weight loss/faltering growth, nutritional deficiencies, nutritional supplement dependence; DSM-5 Criteria A1-A3) may be relatively straightforward, assessing marked interference with psychosocial functioning (DSM-5 Criterion A4) can be more challenging, especially in young children. Specifically, it may be difficult to distinguish the negative impacts on parents and family life from the negative impact on the child, which is required to meet Criterion A4. Well-validated ARFID screening tools assessing these criteria, particularly tools based on parent-reports, are currently lacking (Dinkler & Bryant-Waugh, 2021).

To address this gap, we developed the ARFID-Brief Screener, a parent-reported questionnaire closely aligned with DSM-5 criteria. When tested among 4- to 7-year-old Japanese children, 1.3% screened positive for ARFID, with an approximately equal distribution between boys and girls (Dinkler et al., 2022a), consistent with other prevalence and sex distribution estimates (Sanchez-Cerezo et al., 2023). We also demonstrated the screener’s convergent validity with anthropometric measures and restrictive food intake behaviors, offering initial support for this tool (Dinkler et al., 2022a). However, its diagnostic validity against clinical ARFID diagnoses remains unclear.

The differentiation of normative selective eating from ARFID could further benefit from a better understanding of early risk factors and clinical characteristics in preschoolers with ARFID. For example, a strong overrepresentation of neurodevelopmental conditions such as autism, attention deficit hyperactivity disorder (ADHD), and intellectual disability in ARFID is well established (Dinkler et al., 2022b; Nyholmer et al., 2025; Sanchez-Cerezo et al., 2023; Wronski et al., 2025). Since these conditions are typically characterized by early onset, their presence may indicate an increased likelihood of ARFID when evaluating persistent eating problems in preschoolers, although this area remains underexplored in young children (Dinkler et al., 2022b).

This study aimed to enhance the understanding of ARFID presentation in preschool children, evaluate a screening tool for its identification, and thereby improve detection in this population via the following specific aims: (1) identify children with ARFID in a population of Swedish preschoolers attending preventative health care visits by using the ARFID-Brief Screener in combination with a diagnostic interview for ARFID; (2) estimate the point prevalence of ARFID in this population; (3) describe the clinical characteristics of preschool children with ARFID, including anthropometrics, ARFID severity and profiles, and co-occurring neurodevelopmental conditions; and (4) provide further data on the psychometric characteristics of the ARFID-Brief Screener by evaluating its agreement with a diagnostic interview.

## 2 Methods

### 2.1 Participants

#### 2.2.1 Study population

The study population consisted of Swedish children born between June 2016 and April 2020, visiting one of 21 child health services (CHS) centers in the Fyrbodal region, north of Gothenburg, West Sweden. Between November 2020 and June 2022, parents were invited by CHS staff to participate in the study in connection with routine check-ups at age 2.5 (children born February 2018–April 2020) and age 4 (children born June 2016–June 2018). Follow-up telephone interviews and CHS records were collected until June 2023. Participating CHS staff received one hour of training on ARFID and the study procedures. The study was approved by the Swedish Ethical Review Authority (no. 2020-01284, 2020-03908, 2021-01849).

#### 2.2.2 Dropout and response rate

Of all 37 eligible CHS centers in the Fyrbodal region, 22 (59.5%) agreed to participate, although one CHS center dropped out during data collection. Approximately 5,000 children were expected to attend routine check-ups at the remaining 21 CHS centers between November 2020 and June 2022, with around 70% (n∼3,500) of parents having sufficient Swedish or English skills to participate. Due to the COVID-19 pandemic and other factors, only about 50% of eligible participants (n∼1,750) were invited by CHS staff, and of these, 670 participated (38.3%; see limitations section). After excluding 25 participants due to invalid consent forms or unclear screening results, the final sample included 645 children (**Figure 1**).

**Figure 1.**
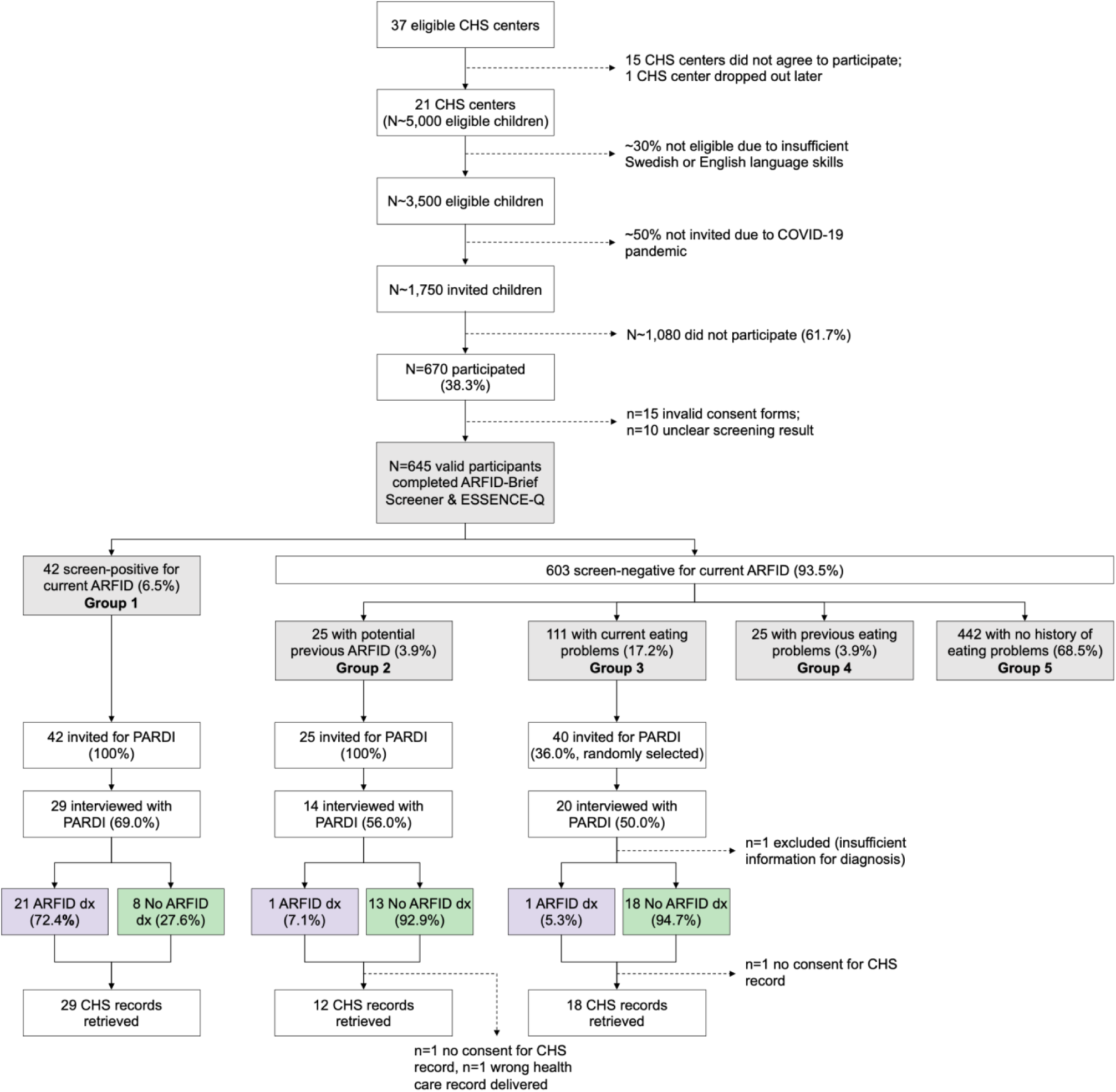
Flowchart showing participation, data availability, screening groups, and ARFID status. Formula used to calculate ARFID point prevalence: [Number of children in group 1 * Proportion with ARFID diagnosis in group 1 + Number of children in group 2 * Proportion with ARFID diagnosis in group 2 + Number of children in group 3 * Proportion with ARFID diagnosis in group 3] / Total sample size. Across age groups: [42 * (21 / 29) + 25 * (1 / 14) + 111 * (1 / 19)] / 645 = 5.9% 2.5-year-olds: [21 * (11 / 15) + 16 * (1 / 10) + 64 * (0 / 11)] / 371 = 4.6% 4-year-olds: [21 * (10 / 14) + 9 * (0 / 4) + 47 * (1 / 8)] / 274 = 7.6% ARFID: avoidant restrictive food intake disorder; CHS: child health services; PARDI: Pica, ARFID, and Rumination Disorder Interview.

### 2.2 Measures

Together with the invitation to the CHS routine visit, CHS staff sent out an invitation to the study, the ARFID-Brief Screener, and a screener for neurodevelopmental problems. Parents/guardians (henceforth referred to as *parents* for readability) were supposed to bring the completed screeners to the visit. If they forgot to bring them to the clinic, they received new forms to fill out at the visit. After the visit, the completed questionnaires were sent to the research team, who then invited parents of eligible children to complete an in-depth diagnostic interview for ARFID by phone.

#### 2.2.1 ARFID–Brief Screener

The ARFID–Brief Screener is a short, parent-reported tool designed to screen for ARFID in children aged 2 to 17, assessing both current and lifetime perspectives (i.e., current and previous ARFID symptoms) (Dinkler et al., 2022a). Items and diagnostic algorithm align closely with the DSM-5 diagnostic criteria for ARFID (**Table S1**). Screening status was determined as outlined in Table 1. The main differentiation was made between screening positive versus negative for current ARFID, but negative screens were further subdivided for validation purposes, which was possible due to the assessment of current and previous ARFID symptoms (**Figure 1**). In prior research, the ARFID–Brief Screener v1 demonstrated satisfactory convergent validity with problems related to mealtime behaviors, nutritional intake, selective eating, and satiety responsiveness, as well as with shorter height and lower body mass index (BMI) (Dinkler et al., 2022a). This study used a revised version of the screener (ARFID–Brief Screener *v2*, **Table S1;** for modifications see Table S1 in (Dinkler et al., 2022a)).

**Table 1.**
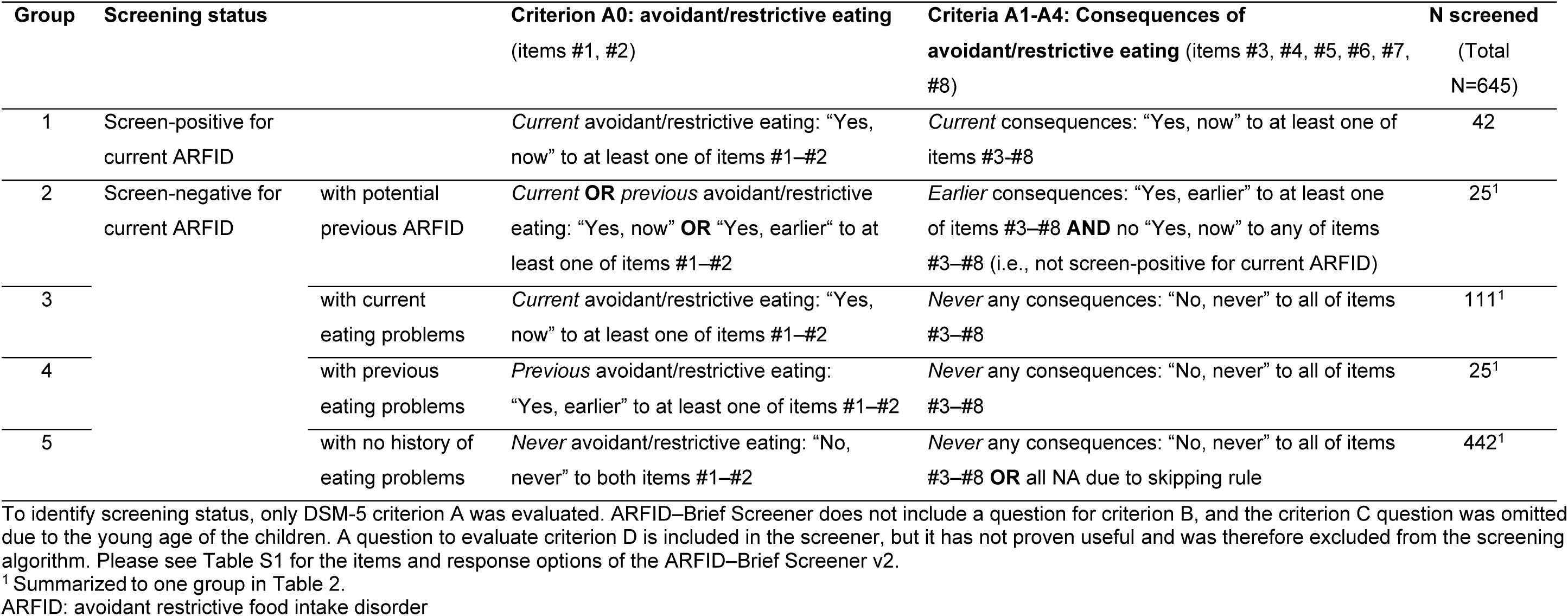
Identification of screening status according to ARFID–Brief Screener v2 and available data by screening status.

| Group | Screening status |  | Criterion A0: avoidant/restrictive eating<br>(items #1, #2) | Criteria A1-A4: Consequences of<br>avoidant/restrictive eating (items #3, #4, #5, #6, #7, #8) | N screened<br>(Total<br>N=645) |
| --- | --- | --- | --- | --- | --- |
| 1 | Screen-positive for current ARFID |  | *Current* avoidant/restrictive eating: “Yes, now” to at least one of items #1–#2 | *Current* consequences: “Yes, now” to at least one of items #3–#8 | 42 |
| 2 | Screen-negative for current ARFID | with potential previous ARFID | *Current* **OR** *previous* avoidant/restrictive eating: “Yes, now” **OR** “Yes, earlier“ to at least one of items #1–#2 | *Earlier* consequences: “Yes, earlier” to at least one of items #3–#8 **AND** no “Yes, now” to any of items #3–#8 (i.e., not screen-positive for current ARFID) | 25<sup>1</sup> |
| 3 |  | with current eating problems | *Current* avoidant/restrictive eating: “Yes, now” to at least one of items #1–#2 | *Never* any consequences: “No, never” to all of items #3–#8 | 111<sup>1</sup> |
| 4 |  | with previous eating problems | *Previous* avoidant/restrictive eating: “Yes, earlier” to at least one of items #1–#2 | *Never* any consequences: “No, never” to all of items #3–#8 | 25<sup>1</sup> |
| 5 |  | with no history of eating problems | *Never* avoidant/restrictive eating: “No, never” to both items #1–#2 | *Never* any consequences: “No, never” to all of items #3–#8 **OR** all NA due to skipping rule | 442<sup>1</sup> |

<sup>1</sup> Summarized to one group in Table 2.
ARFID: avoidant restrictive food intake disorder

#### 2.2.2 Diagnostic interview for ARFID

Parents from three screening groups were invited to a telephone interview using the parent-report version of the Pica, ARFID, and Rumination Disorder Interview (PARDI) (Bryant-Waugh et al., 2019): (1) all children screening positive for current ARFID; (2) all children screening negative for current ARFID but with potential previous ARFID; and (3) randomly selected screen-negative children with current eating problems not meeting ARFID criteria (cf. groups 1-3 in **Figure 1 & Table 1**). The PARDI is a semi-structured diagnostic interview for ARFID employing an established diagnostic algoritm concept (e.g., also embedded in the Eating Disorders Examination [EDE] (Fairburn et al., 2014)). In addition to a diagnostic decision for ARFID, the PARDI provides a severity score and three profile scores (Sensory, Low interest, Concern). A validation study in 9–23-year-olds supported this 3-profile structure and demonstrated excellent discrimination between clinical and non-clinical ARFID cases for two of the three profile scores (Low interest: cutoff 1.1, 83.1% correctly classified cases; Sensory: cutoff 0.6; 84.4% correctly classified cases) (Cooper-Vince et al., 2022). However, the applicability of theses clinical cut-offs in preschool children is yet unclear as no published studies have evaluated the discriminatory ability of the PARDI in this population. The questions screening for weight and shape concerns— which could indicate another eating disorder and thus rule out an ARFID diagnosis (DSM-5 Criterion C)—were omitted due to the children’s young age. The PARDI also includes a health checklist to contextualize eating behaviors and identify potential exclusion criteria for an ARFID diagnosis (DSM-5 Criterion D).

The PARDI can be used by clinicians and researchers from diverse professional backgrounds; however, training is essential to ensure the interviewer fully understands the standardized items and can paraphrase them effectively to gather accurate information. To achieve a precise understanding of what each PARDI item assesses, our team received training delivered in English by RBW. The PARDI was subsequently translated into Swedish by our team, which included both native and non-native speakers with excellent English proficiency. Any ambiguities in the translation were thoroughly discussed and resolved, and as a result, a 1:1 back-translation was not deemed necessary. Two interviewers, an MSc-level psychologist and a medical student (KB and HH), conducted the PARDI interviews after attending training. To assess interrater reliability, ten participants were rated by both interviewers, with 100% agreement on the diagnostic decision and on whether criteria A2, A3, and A4 were met; agreement for Criterion A1 was 90%. Intraclass correlation coefficients (ICC) for the profile and severity scores ranged from 0.95 (95% confidence interval [CI] 0.83-0.99) to 0.97 (95% CI 0.89-0.99), indicating high interrater agreement.

#### 2.2.3 Clinical characteristics

Clinical characteristics of ARFID presentation and neurodevelopmental conditions were compared between children with and without an ARFID diagnosis according to the PARDI. Data were extracted from the following sources: (1) ARFID-Brief Screener (parent-report, described in detail above), (2) PARDI (interviewer rating of parent-report, described in detail above), (3) CHS records, and (4) a parent-reported screener for neurodevelopmental conditions (ESSENCE-Q). **Table 2** presents the clinical characteristics extracted from each source.

**Table 2.** Data sources for clinical characteristics in 62 children with available PARDI data.

| Clinical characteristic | ARFID-Brief Screener | PARDI | ESSENCE-Q | CHS records |
| --- | --- | --- | --- | --- |
| Type of information | Parent-report | Interviewer rating of parent-report | Parent-report | Clinical records |
| Available for | n=62 | n=62 | n=62 | n=59 |
| ARFID presentation | X | X | --- | X (growth charts) |
| Anthropometrics | X | X | --- | X (growth charts) |
| ARFID criteria | X | X | --- | -- |
| ARFID profiles | X | X | --- | -- |
| ARFID severity | --- | X | --- | -- |
| Neurodevelopmental problems | --- | X (health checklist) | X | X (diagnoses, result from structured speech and language test) |

ARFID: avoidant restrictive food intake disorder; CHS: child health services; ESSENCE-Q: Early Symptomatic Syndromes Eliciting Neurodevelopmental Clinical Examinations Questionnaire; PARDI: Pica, ARFID, and Rumination Disorder Interview.

### CHS records

CHS records were collected for children interviewed with the PARDI (groups 1-3, **Figure 1**). We extracted information on suspected or diagnosed neurodevelopmental conditions and results from the structured speech and language test, which is routinely conducted during the 2.5-year check-up with ∼97% coverage (Västa Götalandsregionen, 2021). Previous research indicates that children with delayed language development are at significantly increased risk of neurodevelopmental conditions such as autism and ADHD (Miniscalco et al., 2006).

### ESSENCE-Q

Along with the ARFID–Brief Screener, all participants completed the 12-item *Early Symptomatic Syndromes Eliciting Neurodevelopmental Clinical Examinations Questionnaire* (ESSENCE-Q; Hatakenaka et al., 2016). ESSENCE-Q screens for early neurodevelopmental problems that might indicate the presence of neurodevelopmental conditions and the need for further clinical examination. The screener is used in clinical and research settings across multiple countries (Landgren et al., 2022). Responses are rated as *“Yes” (2), “Maybe/a little” (1)* or *“No” (0),* yielding a total score ranging from 0 to 24. In the current study, two items were omitted from the total score. The item *Feeding problems* was excluded because it was endorsed for all children with ARFID (“Yes”: 91.3%, “Maybe/a little”: 8.7%; **Table S3**). The item *Sensory reactions* (e.g., touch, sound, light, smell, taste, heat, cold, pain) was excluded because sensory aversions to food characteristics are an inherent and common symptom of ARFID, even though this item was only reported in a subset of children with ARFID (“Yes”: 13.6%, “Maybe/a little”: 13.6%; **Table S3)**. Due to these modifications to the total ESSENCE-Q score, we refrained from using previously suggested cutoff values (Landgren et al., 2022) in the current study.

### 2.3 Statistical Analyses

Analyses were conducted in R version 4.2.3 (R Core Team, 2022). The significance level was set at 0.05. Interrater reliability for the PARDI was evaluated using ICCs within the *irr* package (Gamer et al., 2019). Group differences were tested using Wilcoxon rank sum test for continuous variables and Pearson’s Chi-squared test (if all expected cell counts ≥5) or Fisher’s exact test (if any expected cell count <5) for categorical variables, using the *gtsummary* package (Sjoberg et al., 2021). Due to small group sizes and the main goal of describing clinical characteristics in the ARFID group (focusing on absolute values), we did not include covariates or correct for multiple testing. Additionally, the number of females in the ARFID group was too small to include sex as a covariate.

#### 2.3.1 Estimation of ARFID point prevalence

Point prevalence of ARFID was estimated across age groups and separately by age group (i.e., in 2.5-year-olds and 4-year-olds; see **Figure 1** for calculation formula). Estimations assumed (1) the same proportion of children with ARFID among those interviewed with the PARDI as among those who declined the interview, and (2) that there were no children with ARFID among those screen-negative with previous eating problems (group 4) and those screen-negative with no history of eating problems (group 5; none of the children in these groups were followed up with the PARDI).

#### 2.3.2 Agreement between ARFID-Brief Screener and PARDI

To test the agreement between the screening result from the ARFID-Brief Screener and the diagnostic status obtained in PARDI, we assessed statistical validity using the *epiR* package (Stevenson et al., 2022). We report sensitivity, specificity, PPV, NPV, and accuracy. Due to the imbalanced class distribution (more true negatives [no ARFID diagnosis] than true positives [ARFID diagnosis]), we also report the F1 Score, which combines precision (PPV) and recall (sensitivity) into a single metric, providing a balanced measure of diagnostic accuracy. Like other diagnostic metrics, the F1 score ranges from 0 (PPV=0 or sensitivity=0) to 1 (PPV=1 and sensitivity=1). The overall test of statistical validity was calculated across age groups, including both children with potential previous ARFID (group 2) and children with current eating problems not meeting ARFID criteria (group 3) into the group with a negative screening result. Supplemental statistical validity tests were conducted separately by age group (i.e., in 2.5-year-olds and 4-year-olds) and separately for groups 2 and 3.

## 3 Results

Among the 645 children with valid ARFID-Brief Screener results, the sex distribution was nearly equal (50.5% male), with slightly more participants recruited during the 2.5-year check-up (57.5%; **Table 3**). The average age at screening was 38 months, ranging from 20 to 59 months (**Table 1**). In rare cases, the visits were conducted up to 11 months before or after the child turned 2.5/4 years. Most respondents were mothers (83.8%). Current avoidant/restrictive eating problems (DSM-5 Criterion A0) were reported in 25.3% of all children. Children screening positive for current ARFID (n=42, 6.5%) had significantly lower BMIs and were more likely to have parents born outside Sweden. No differences were observed in parental education levels. Sample characteristics for all screening groups are provided in **Table S2**. A total of 62 PARDI interviews were conducted (**Figure 1**). In 59 of these, the respondent was the same person who filled in the ARFID-Brief Screener (of which 56 were mothers). Both parents were present in 4 of the 62 interviews.

**Table 3.** Sample characteristics overall and comparing children screening positive for current ARFID to children screening negative for current ARFID (all other screening groups summarized)

| Characteristic | Overall<br>N = 645 | Screen-positive for<br>current ARFID<br>N = 42 | Screen-negative for<br>current ARFID<br>N = 603 | p-value<sup>1</sup> |
| --- | --- | --- | --- | --- |
| **Sex** |  |  |  | 0.942 |
| Female | 319 (49.5%) | 21 (50.0%) | 298 (49.4%) |  |
| Male | 326 (50.5%) | 21 (50.0%) | 305 (50.6%) |  |
| **Age at screen (months)** | 38.0 (9.0), 20.4-59.1 | 39.2 (9.6), 24.4-51.4 | 38.0 (9.0), 20.4-59.1 | 0.552 |
| **Routine check-up** |  |  |  | 0.308 |
| 2.5 years | 371 (57.5%) | 21 (50.0%) | 350 (58.0%) |  |
| 4 years | 274 (42.5%) | 21 (50.0%) | 253 (42.0%) |  |
| **Current BMI** | 16.4 (1.5), 12.7-24.4 | 15.3 (1.4), 12.7-18.8 | 16.5 (1.4), 13.4-24.4 | **<0.001** |
| **Respondent** |  |  |  | 0.454 |
| Mother | 538 (83.8%) | 36 (87.8%) | 502 (83.5%) |  |
| Father | 93 (14.5%) | 4 (9.8%) | 89 (14.8%) |  |
| Mother & Father | 11 (1.7%) | 1 (2.4%) | 10 (1.7%) |  |
| **Mother born in Sweden** | 521 (80.8%) | 27 (64.3%) | 494 (81.9%) | **0.005** |
| **Father born in Sweden** | 511 (79.8%) | 27 (64.3%) | 484 (80.9%) | **0.009** |
| **Mother's education<sup>2</sup>** |  |  |  | 0.360 |
| Less than compulsory school (<9y) | 11 (1.7%) | 1 (2.4%) | 10 (1.7%) |  |
| Compulsory school (9y) | 32 (5.0%) | 3 (7.1%) | 29 (4.8%) |  |
| High school (12y) | 184 (28.7%) | 16 (38.1%) | 168 (28.0%) |  |
| Further education (13-14y) | 145 (22.6%) | 9 (21.4%) | 136 (22.7%) |  |
| University/college degree (15+y) | 269 (42.0%) | 13 (31.0%) | 256 (42.7%) |  |
| **Father's education<sup>2</sup>** |  |  |  | 0.172 |
| Less than compulsory school (<9y) | 14 (2.2%) | 2 (4.8%) | 12 (2.0%) |  |
| Compulsory school (9y) | 45 (7.1%) | 5 (11.9%) | 40 (6.8%) |  |
| High school (12y) | 286 (45.4%) | 14 (33.3%) | 272 (46.3%) |  |
| Further education (13-14y) | 111 (17.6%) | 10 (23.8%) | 101 (17.2%) |  |
| University/college degree (15+y) | 174 (27.6%) | 11 (26.2%) | 163 (27.7%) |  |
| **DSM-5 Criterion A0** |  |  |  | ---- |
| Yes, currently | 163 (25.3%) | 42 (100.0%)<sup>3</sup> | 121 (20.1%) |  |
| Yes, earlier | 40 (6.2%) | 0 (0.0%)<sup>3</sup> | 40 (6.6%) |  |
| No, never | 442 (68.5%) | 0 (0.0%)<sup>3</sup> | 442 (73.3%) |  |

Sample characteristics for each screening group separately are presented in Table S2.
<sup>1</sup> Categorical variables: n (%); Pearson's Chi-squared test (all expected cell counts $\geq 5$ ) or Fisher's exact test (any expected cell count $< 5$ ). Continuous variables: M (SD), Min-Max; Wilcoxon rank sum test. <sup>2</sup> "y" indicates total number of total school years. <sup>3</sup> By definition: meeting criterion A0 (avoidant/restrictive eating) was required for this screening group.

### 3.1 ARFID point prevalence

The estimated overall point prevalence across age groups was 5.9% (see **Figure 1** for calculation formula). Point prevalence was 4.6% in 2.5-year-olds and 7.6% in 4-year-olds; this difference was not statistically significant, *χ*^2^(1)=2.11, *p*=.147.

### 3.2 Clinical characteristics

Children diagnosed with ARFID in the PARDI (n=39) were similar to those without an ARFID diagnosis (n=23) in terms of age at screening, age at PARDI assessment, and expected height (“control variables”, **Table 4**). The two groups also showed no difference in the age of onset of eating problems. Although not statistically significant (p=.066), the proportion of males with ARFID (62.5%) was higher than in the non-ARFID group (41.0%). No significant differences were observed in anthropometric variables, though there was a trend towards lower BMI in children with ARFID. Among children with ARFID, 26.1% were underweight (degree 1 or 2), 65.2% had normal weight, and 8.7% had overweight.

**Table 4.** Demographic and clinical characteristics including ARFID presentation and neurodevelopmental problems in children with versus without ARFID diagnosis (dx) in the PARDI.

| Characteristic | No ARFID dx in PARDI<br>N = 39 | ARFID dx in PARDI<br>N = 23 | p-value<sup>1</sup> |
| --- | --- | --- | --- |
| **Demographics** |  |  |  |
| Sex |  |  | 0.066 |
| Female | 23 (59.0%) | 8 (34.8%) |  |
| Male | 16 (41.0%) | 15 (65.2%) |  |
| Routine check-up |  |  | 0.470 |
| 2.5 years | 24 (61.5%) | 12 (52.2%) |  |
| 4 years | 15 (38.5%) | 11 (47.8%) |  |
| Age (Screen), months | 37.1 (9.0), 28.4-51.1 | 38.7 (9.5), 24.4-50.7 | 0.502 |
| Age (PARDI), months | 40.2 (8.9), 29.8-56.2 | 41.4 (10.0), 25.8-60.3 | 0.810 |
| Δ Time PARDI – Screen, months | 3.1 (1.8), 0.9-8.2 | 2.7 (1.9), 0.8-9.9 | 0.197 |
| Age at onset of eating problems, months | 12.8 (10.2), 0.0-40.0 | 12.4 (9.7), 0.0-36.0 | 0.892 |
| Mother born in Sweden | 34 (87.2%) | 17 (73.9%) | 0.302 |
| Father born in Sweden | 30 (76.9%) | 17 (73.9%) | 0.789 |
| Mother's education |  |  | 0.362 |
| Less than compulsory school (<9y) | 1 (2.6%) | 1 (4.3%) |  |
| Compulsory school (9y) | 0 (0.0%) | 2 (8.7%) |  |
| High school (12y) | 13 (33.3%) | 8 (34.8%) |  |
| Further education (13-14y) | 12 (30.8%) | 4 (17.4%) |  |
| University/college degree (15+y) | 13 (33.3%) | 8 (34.8%) |  |
| Father's education |  |  | 0.263 |
| Less than compulsory school (<9y) | 1 (2.6%) | 0 (0.0%) |  |
| Compulsory school (9y) | 2 (5.3%) | 3 (13.0%) |  |
| High school (12y) | 23 (60.5%) | 9 (39.1%) |  |
| Further education (13-14y) | 4 (10.5%) | 6 (26.1%) |  |
| University/college degree (15+y) | 8 (21.1%) | 5 (21.7%) |  |
| **Anthropometrics** |  |  |  |
| Expected height in SD (PARDI) | 0.0 (0.8), -1.7-2.0 | -0.2 (1.0), -2.5-2.2 | 0.323 |
| Height in SD (PARDI) | -0.3 (1.1), -4.0-2.0 | -0.2 (1.0), -2.0-1.4 | 0.965 |
| Weight in SD (PARDI) | -0.5 (1.1), -2.5-1.5 | -0.6 (1.2), -3.4-1.8 | 0.692 |
| BMI (PARDI) | 15.9 (1.4), 13.7-19.4 | 15.7 (1.4), 12.7-18.5 | 0.466 |
| BMI (Screen) | 15.9 (1.4), 13.8-19.4 | 15.7 (1.4), 12.7-18.7 | 0.562 |
| Δ BMI PARDI – Screen | 0.0 (0.2), -1.0-0.4 | 0.0 (0.1), -0.2-0.5 | 0.903 |
| BMI in SD (PARDI) | -0.3 (1.2), -3.0-2.0 | -0.7 (1.1), -3.0-1.2 | 0.220 |
| BMI in IOTF category (PARDI) |  |  | 0.864 |
| Underweight degree 2 (BMI 16.0 - <17.0)<sup>2</sup> | 3 (7.7%) | 2 (8.7%) |  |
| Underweight degree 1 (BMI 17.0 - <18.5)<sup>2</sup> | 4 (10.3%) | 4 (17.4%) |  |
| Normal weight (BMI 18.5 - <25.0)<sup>2</sup> | 29 (74.4%) | 15 (65.2%) |  |
| Overweight (BMI 25.0 - <30.0)<sup>2</sup> | 3 (7.7%) | 2 (8.7%) |  |
| **ARFID criteria (Screener)** |  |  |  |
| Criterion A1: Yes, currently | 6 (15.4%) | 7 (30.4%) | 0.203 |
| Criterion A2: Yes, currently | 0 (0.0%) | 3 (13.0%) | 0.047 |
| Criterion A3: Yes, currently | 1 (2.6%) | 6 (26.1%) | 0.009 |
| Criterion A4: Yes, currently | 3 (8.1%) | 15 (65.2%) | <.001 |
| **ARFID criteria (PARDI)** |  |  |  |
| Criterion A0 met | 22 (56.4%) | 23 (100.0%)<sup>3</sup> | ---- |
| Criterion A1 met | 0 (0.0%)<sup>3</sup> | 3 (13.0%) | ---- |
| Criterion A2 met | 0 (0.0%)<sup>3</sup> | 1 (4.3%) | ---- |
| Criterion A3 met | 0 (0.0%)<sup>3</sup> | 6 (26.1%) | ---- |
| Criterion A4 met | 0 (0.0%)<sup>3</sup> | 20 (87.0%) | ---- |
| Stressful mealtimes (Q42) | 0 (0.0%)<sup>3</sup> | 14 (70.0%) | ---- |
| Social difficulties (Q47) | 0 (0.0%)<sup>3</sup> | 6 (30.0%) | ---- |
| Daily functioning difficulties (Q48) | 0 (0.0%)<sup>3</sup> | 12 (60.0%) | ---- |
| # of criteria met (of A1-A4) |  |  | ---- |
| 0 | 39 (100.0%)<sup>3</sup> | 0 (0.0%)<sup>3</sup> | ---- |
| 1 | 0 (0.0%)<sup>3</sup> | 17 (73.9%) | ---- |
| 2 | 0 (0.0%)<sup>3</sup> | 5 (21.7%) | ---- |
| 3 | 0 (0.0%)<sup>3</sup> | 1 (4.3%) | ---- |
| >1 criterion met | 0 (0.0%)<sup>3</sup> | 6 (26.1%) | ---- |
| Only criterion A4 met (A1-A4) | 0 (0.0%)<sup>3</sup> | 15 (65.2%) | ---- |
| **ARFID profiles (PARDI)** |  |  |  |
| Severity score (TR 0-6) | 1.1 (0.7), 0.2-3.0 | 2.8 (0.6), 1.9-4.3 | <0.001 |
| Sensory profile score (TR 0-6) | 1.0 (0.7), 0.0-3.2 | 2.3 (1.2), 0.7-5.1 | <0.001 |
| Low interest profile score (TR 0-6) | 1.6 (0.7), 0.4-3.1 | 2.8 (0.7), 1.6-4.0 | <0.001 |
| Concern profile score (TR 0-6) | 0.0 (0.1), 0.0-0.4 | 0.2 (0.3), 0.0-1.4 | 0.035 |
| # Sensory profile items ≥4 (TR 0-10) | 1.3 (1.3), 0.0-5.0 | 4.0 (2.3), 1.0-8.0 | <0.001 |
| # Low interest profile items ≥4 (TR 0-10) | 1.7 (1.6), 0.0-5.0 | 4.9 (2.1), 1.0-8.0 | <0.001 |
| # Concern profile items ≥4 (TR 0-10) | 0.0 (0.2), 0.0-1.0 | 0.1 (0.5), 0.0-2.0 | 0.281 |
| Sensory profile score ≥0.6 | 30 (76.9%) | 23 (100.0%) | 0.020 |
| Low interest profile score ≥1.1 | 31 (79.5%) | 23 (100.0%) | 0.021 |
| **Neurodevelopmental problems** |  |  |  |
| Modified ESSENCE-Q score (TR 0-20) | 2.0 (2.9), 0-12 | 3.6 (4.4), 0-14 | 0.166 |
| Failed language screen at 2.5 years | 5 (13.5%) | 9 (39.1%) | 0.023 |
| Diagnosed or suspected autism | ---- | 5 (21.7%)<sup>5</sup> | ---- |
| Diagnosed or suspected ADHD | ---- | 4 (17.4%)<sup>6</sup> | ---- |
| Diagnosed intellectual disability | ---- | 0 (0.0%) | ---- |

<sup>1</sup> Categorical variables: n (%), Pearson's Chi-squared test (all expected cell counts ≥5) or Fisher's exact test (any expected cell count <5). Continuous variables: M (SD), Min-Max; Wilcoxon rank sum test. <sup>2</sup> Adult BMI. <sup>3</sup> By definition: meeting criterion A0 and at least one of criteria A1-A4 was required for ARFID dx. <sup>4</sup> Not reported as we did not obtain sufficient information for all children in this group. <sup>5</sup> Four children also failed the language screen at 2.5 years. <sup>6</sup> One child also failed the language screen at 2.5 years. ARFID: avoidant restrictive food intake disorder; PARDI: Pica, ARFID, and Rumination Disorder Interview; TR: Theoretical range.

All children who met Criterion A also met Criterion D, that is, their eating problems were not attributable to, or better explained by, another medical or mental condition. Most children with ARFID (n=15, 65.2%) met criteria solely through Criterion A4 (marked interference with psychosocial functioning), while only 13.0% showed a negative impact on weight or height (Criterion A1), consistent with the non-significant anthropometric differences. Notably, 26.1% of children with ARFID met two or more diagnostic criteria (A1 to A4). PARDI severity and profile scores were significantly higher in children with ARFID compared to those without (**Table 4**, **Figure 2**). All children with ARFID scored above cutoff on both Sensory and Low interest profiles.

**Figure 2.**
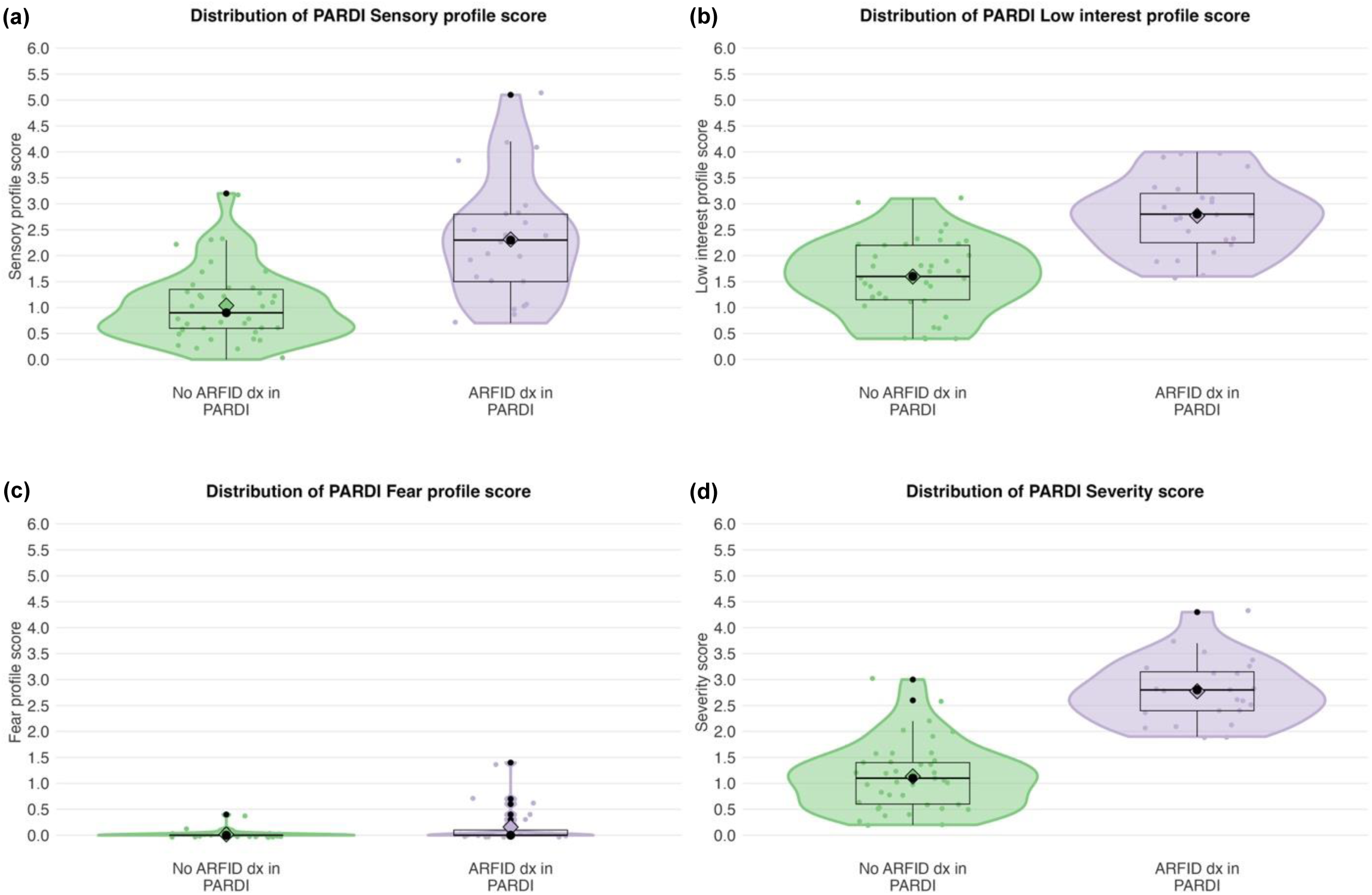
Distribution of PARDI profile scores (a-c) and PARDI severity score (d). ARFID: avoidant restrictive food intake disorder; PARDI: Pica, ARFID, and Rumination Disorder Interview.

The mean difference in the modified ESSENCE-Q score was not statistically significant (M=3.6 vs. M=2.0; *p*=.166). Among the individual ESSENCE-Q items, only *Feeding problems* and *Sensory reactions* were significantly more frequent in children with ARFID (*p*=.039; **Table S3**). However, children with ARFID showed a general tendency toward higher prevalence across all ESSENCE-Q items (except *“Funny spells”/absences,* **Table S3**) and they were nearly three times more likely to fail the language screen at 2.5 years (39.1% vs. 13.5%, *p*=.023). The modified ESSENCE-Q score correlated significantly with the PARDI scores for Severity (*r*=.47, *p*=.023), Sensory profile (*r*=.54, *p*=.008), and Concern profile (*r*=.54, *p*=.008), but not with the Low interest profile (*r*=-.08, *p*=.728).

### 3.3 Agreement between ARFID-Brief Screener and PARDI

Results from the overall test of statistical validity are shown in **Table 5**. Among 29 children screening positive for current ARFID, a diagnosis was confirmed in 21 (PPV 72%). Of the 33 screen-negative children interviewed with the PARDI, the absence of ARFID was confirmed for 31 children (NPV 94%). Sensitivity, specificity, overall accuracy, and F1 score were 91%, 79%, 84%, and 81%, respectively. Classification accuracy was similar between different definitions of a negative screening result (those with potential previous ARFID [group 2] vs. those with current eating problems not meeting ARFID criteria [group 3]) and between age groups, though confidence intervals were large due to small sample sizes (**Table S4**).

**Table 5.** Agreement between ARFID-Brief Screener (Screen +/-) and clinical interview (PARDI +/-) across age groups.

|  | **PARDI +** | **PARDI -** |
| --- | --- | --- |
| **Screen +** | 21 | 8 |
| **Screen -** | 2 | 31 |

*Statistical tests*
| **Statistic** | **Estimate (95%CI)** |
| --- | --- |
| Sensitivity | 0.91 (0.72-0.99) |
| Specificity | 0.79 (0.64-0.91) |
| Positive predictive value | 0.72 (0.53-0.87) |
| Negative predictive value | 0.94 (0.80-0.99) |
| Accuracy | 0.84 (0.72-0.92) |
| F1 Score<sup>1</sup> | 0.81 (0.76-0.94) |

<sup>1</sup> Bootstrapped confidence intervals
Screen +: screen-positive for current ARFID (group 1)
Screen -: screen-negative for current ARFID but with potential previous ARFID (group 2) or current eating problems (group 3)
PARDI +: ARFID diagnosis in PARDI
PARDI -: no ARFID diagnosis in PARDI
ARFID: avoidant restrictive food intake disorder; PARDI: Pica, ARFID, and Rumination Disorder Interview.

### 3.4 Analysis of sample representativeness

To address concerns about sample representativeness given the relatively low response rate (38.3%), we compared demographic variables (parents born in Sweden vs. foreign-born; parental education levels) with data from Statistics Sweden (www.scb.se). For the year 2021, when most of our data collection occurred, we filtered statistics to include individuals within a reasonable parental age range (20-49 years, given the age of the children in this study) who resided in the nine municipalities where the CHS centers were located. Comparing to this population, parents in our sample were slightly more likely to be born in Sweden (81.0% vs. 72.8%) and had somewhat higher levels of education (e.g., 13-14 years of education: 20.1% vs. 15.3%, 15+ years of education: 34.9% vs. 26.4%). However, it was not possible to determine whether individuals in the comparison population were parents.

## 4 Discussion

This study provides a first estimate of ARFID point prevalence in a community-based sample of preschool children and a detailed characterization of clinical features in preschool children with ARFID. We also evaluated the psychometric performance of the ARFID-Brief Screener in this population.

### Prevalence of ARFID

The overall point prevalence of ARFID was estimated at 5.9%, with higher rates observed in 4 - year-olds (7.6%) compared to 2.5-year-olds (4.6%). These estimates are difficult to compare with previous findings in non-clinical samples, as reported estimates have varied widely, particularly for parent-reported ARFID symptoms, which range from 1.3% to 15.5% in 4- to 12-year-olds across studies conducted in Japan, Sweden, the Netherlands, Israel, and Portugal (Dinkler et al., 2023; Dinkler et al., 2022a; Gonçalves et al., 2018; Iron-Segev et al., 2023; Sader et al., 2023). Furthermore, no previous studies have specifically targeted preschool children (Sanchez-Cerezo et al., 2023). Differences in age, cultural contexts, and assessment methods likely contribute to this variability. The only other study using the ARFID-Brief Screener was conducted in a slightly older sample of Japanese children (ages 4–7) and reported a much lower prevalence (1.3%) than observed in the current study (Dinkler et al., 2022a). While this discrepancy may be partly explained by age differences, it could also reflect cultural differences in the perception and reporting of eating problems. Based on our experience, discussing such concerns may be less culturally accepted in Japan than in Sweden, which may have contributed to the lower prevalence reported in the Japanese sample.

In addition, our estimates likely overestimate the true prevalence for several reasons. First, they assume that the proportion of true ARFID cases among those interviewed with the PARDI is similar to that among those who declined participation in the interview. However, this assumption may not hold: some parents may have declined because they felt their child did not have significant problems, potentially leading to an overestimation of PPV and prevalence. Conversely, others may have been unable to participate due to their child’s severe problems, which could result in an underestimation of PPV and prevalence. Additionally, early in the study, some screening sites unintentionally prioritized recruiting children with known eating problems, possibly leading to an overrepresentation of children with eating difficulties in our sample. Lastly, it was not possible to blind interviewers to screening status, which may have introduced rater bias and contributed to an overestimation of both prevalence and statistical validity.

### Clinical presentation of ARFID

In our non-clinical sample, Criterion A4 (marked interference with psychosocial functioning) was the most frequently met criterion for ARFID diagnosis (87.0%), with 65.2% of children meeting this criterion alone, without having notable impacts on weight, height, or nutrition. In contrast, children with ARFID in our sample did not have significantly lower BMIs than those without ARFID and only 13.0% met Criterion A1 (weight loss/faltering growth). This finding is line with previous research showing that older children are more likely than younger children to meet the nutrition and weight-related ARFID criteria (Katzman et al., 2021; Zickgraf, Murray, et al., 2019). This suggests that psychosocial impairment may be an early marker of ARFID, while nutritional and weight-related issues may emerge later as parental control decreases and energy demands increase during puberty (Zickgraf, Murray, et al., 2019). Additionally, the prevalence of Criterion A2 (nutritional deficiencies) was notably low at 4.3%. While this rate is consistent with our previous study in the Japanese child population (4.1%) (Dinkler et al., 2022a), significantly higher rates have been reported in clinical samples (Katzman et al., 2021; Watts et al., 2023). This discrepancy likely reflects the community-based nature of our sample and the potential underdiagnosis of nutritional deficiencies in non-clinical settings due to insufficient testing.

Nevertheless, the rate of children meeting only Criterion A4 (psychosocial impairment) was high (65.2%), though comparisons with previous studies were not possible due to the lack of research on the same age group. In this study, we paid special attention to the risk of overdiagosing Criterion A4 due to the challenge of distinguishing the child’s impairment from parental or familial stress caused by the child’s eating behavior. The impact of a child’s eating behavior on family dynamics and parental wellbeing may be particularly pronounced in young children, increasing the risk of overdiagnosing Criterion A4 in preschool populations. Currently, no clear guidelines exist on how best to evaluate Criterion A4, and prevalence estimates can vary significantly based on definitions applied (Harshman et al., 2021). To address this, we took specific care in evaluating A4, following the guidelines provided by the PARDI. We considered the Criterion A4 met if the child experienced significant difficulties, such as stressful mealtimes or challenges in social and daily functioning at preschool. To gauge impairment, we often employed the question "If these accommodations were not in place, would that cause your child difficulties?", as recommended in the PARDI. Commonly reported difficulties included frequent mealtime conflicts, anxiety and tantrums during meals, eating alone, and avoiding eating situations outside the home. Despite this careful evaluation, many cases were borderline, just meeting the threshold for Criterion A4. It is, however, worth noting that even moderate selective eating in preschool children has been associated with significant psychopathology and family impairment, underscoring the need for clinical intervention (Zucker et al., 2015). Early detection and intervention, regardless of the diagnosis, could prevent the progression of eating problems into more severe weight, nutritional, and health outcomes later in life.

When applying the previously suggested cutoff values for the PARDI Sensory and Low interest profiles (Cooper-Vince et al., 2022), all children with ARFID in our sample scored above cutoff for *both* profiles. This supports the notion that these profiles are commonly associated with early onset and are typically present in younger children (Katzman et al., 2021; Sanchez-Cerezo et al., 2024; Zickgraf, Lane-Loney, et al., 2019). Moreover, this finding aligns with previous research showing that children with the Sensory profile or a combined Sensory and Low interest profile typically have BMIs within the normal range (Sanchez-Cerezo et al., 2024), which was also observed in our study. However, the fact that all children with ARFID—and most children without ARFID—scored above cutoff for both profiles indicates that the previously identified cutoff values, established in older age groups, may not be entirely applicable to preschool-aged children. The low prevalence of the Concern profile in this preschool sample—although consistent with other research in child populations (Sanchez-Cerezo et al., 2024)—may also reflect that fear of something bad happening is less easily inferred from behavior at this age than more observable features such as picky eating and low appetite/interest. Young children may not be able to articulate such fears and might instead say “I don’t like it” or “I’m not hungry” to avoid eating something they perceive as frightening.

### Co-occurring neurodevelopmental problems

Prior research has consistently shown that ARFID is associated with an elevated risk of neurodevelopmental conditions (Dinkler et al., 2022b; Sader et al., 2025; Sanchez-Cerezo et al., 2023; Watts et al., 2023; Wronski et al., 2025). Although children with ARFID in our sample showed an overall tendency toward more co-occurring neurodevelopmental problems, no significant differences between those with and without ARFID were found in the ESSENCE-Q total score or individual items—aside from *Feeding problems* and *Sensory reactions,* which are both core features of ARFID. This may be due to small group sizes resulting in limited statistical power, or the fact that the ESSENCE-Q was parent-reported. Indeed, a significantly higher proportion of children with ARFID (39.1% vs. 13.5%) failed the clinically administered structured speech and language test at age 2.5, indicating delayed language development. Furthermore, 21.7% of children with ARFID had suspected or diagnosed autism, and 17.4% had suspected or diagnosed ADHD, based on clinical records. Although corresponding data were not available for the comparison group—preventing formal statistical comparisons—these numbers are clearly higher than what is typically expected in the general child population (Fast et al., 2024). They are also consistent with a recent meta-analysis reporting an average prevalence of autism in individuals with ARFID of 16.3% (95% CI 8.6–28.5%) (Sader et al., 2025), as well as with our own findings in a community-based sample of older Swedish children, where 13.8% had diagnosed autism and 17.5% had diagnosed ADHD (Wronski et al., 2025).

Our findings also revealed that a higher load of neurodevelopmental problems correlated with higher scores on the Sensory profile, which is consistent with previous findings (Kambanis et al., 2020; Watts et al., 2023). In addition, a higher ESSENCE-Q score was also associated with greater ARFID severity, aligning with the notion that eating problems in children with autism and other neurodevelopmental conditions may be more challenging to treat. Although children with more neurodevelopmental problems also tended to score higher on the Concern profile, these scores were generally very low (all <1.5, theoretical range: 0-6). Regarding specific neurodevelopmental problems, ARFID was linked to delayed language development and sensory sensitivity. These factors may serve as early markers of increased ARFID risk and could potentially be better predictors than common early feeding problems, which are part of normative development, thereby aiding in the early detection of ARFID (Dinkler et al., 2022b).

### Psychometric properties of the ARFID-Brief Screener

The ARFID-Brief Screener demonstrated good psychometric properties in this population when compared to the clinical interview PARDI. With a sensitivity of 91%, the screener effectively identified children with ARFID, minimizing false negatives. This is crucial for population-level screening, where the priority is to ensure that nearly all cases are detected for further evaluation. The specificity of 79% indicates reasonable accuracy in identifying children without ARFID, although there was also a notable rate of false positives. However, this trade-off is acceptable in population-level screening, where the priority is minimizing missed cases. The PPV of 72% indicates that most children who screen positive truly have ARFID. Given the low prevalence of ARFID, this PPV is relatively high and underscores the screener’s utility in identifying true cases. The NPV of 94% is particularly reassuring for parents and healthcare providers, as it suggests that children who screen negative are unlikely to have ARFID. The overall accuracy of 84% and F1 score of 81% reflect the screener’s balance between precision and recall, making it a valuable tool for the early ARFID identification in young children. There was no indication that the classification accuracy of the ARFID-Brief Screnner depended on age (2.5- vs. 4-year-olds), but the sample sizes were too small to draw definite conclusions.

### Strengths and limitations

The study has several strengths, including a large community-based sample and the use of a two-step process involving screening and clinical interviews. It contributes to the scarce literature on ARFID in preschool children and provides the first estimate of ARFID point prevalence in this population, aiding in effective planning and allocation of healthcare resources. Additionally, this study addresses the lack of validated parent-reported screeners for ARFID by providing further psychometric data for the ARFID-Brief Screener, suggesting it may be a valuable tool for use in general child healthcare. Here, the high NPV of the ARFID-Brief-Screener is a particular strength.

However, several important limitations should be considered. First, the COVID-19 pandemic negatively impacted data collection and participation rates. Due to COVID-19-related restrictions and infections, many CHS visits were delayed, canceled, or conducted online, limiting parents’ participation in the study. Additionally, CHS centers faced increased demands due to COVID-19 vaccination programs, which took priority. Our study did not have a decidated researcher at each site, and clinical staff struggled to accommodate the additional time required for recruitment and data collection. As a result, only ∼50% of eligible children were invited to the study, and the response rate was relatively low (38.3%). Our analysis of sample representativeness suggests that participanting parents may have been more affluent than the whole eligible population; however, it is unclear to what extent this might have impacted our results, as the association between parental socioeconomic status and ARFID is not well established. In this study, parental education levels did not differ significantly between children with and without an ARFID diagnosis in the PARDI.

Several potential sources of bias that may have contributed to a slight overestimation of both the prevalence figures and the statistical validity of the ARFID-Brief Screener have been discussed in detail above. Additionally, prevalence estimation was based on the assumption that no children with ARFID were present among those screened negative and had either previous eating problems (group 4) or no history of eating problems (group 5), in other words, an NPV of 100% was assumed for these groups. However, it seems highly unlikely that parents would report that neither they nor anyone else perceives any eating difficulties if the child truly meets criteria for ARFID. Therefore, we assessed the NPV and adjusted for potential false negatives in the groups where assuming an NPV below 100% seemed more plausible—specifically, in children who screened negative for ARFID but had a history of potential ARFID (group 2) or showed some current eating problems (group 3).

## Conclusion

In conclusion, our study provides valuable insights into the occurrence and characteristics of ARFID in preschool children, a group previously underrepresented in research. With an estimated point prevalence of 5.9%, ARFID is not uncommon among Swedish preschoolers. While this estimate aligns with prior research, it may be slightly inflated due to selection biases. Importantly,

ARFID appears to be at least as prevalent as several other neurodevelopmental conditions in this age group, highlighting the need for increased clinical awareness.

All children with ARFID in our study exhibited sensory-based avoidance and low interest in eating, whereas concern about aversive consequences was uncommon. Only a minority exhibited nutritional or growth-related concerns, whereas the majority met diagnostic criteria due to psychosocial challenges related to eating. These findings underscore the importance of assessing the broader psychosocial impact of ARFID beyond physical health metrics such as weight and nutritional status and considering its effects on social interactions, daily functioning, and family dynamics.

Co-occurring neurodevelopmental problems appeared relatively common, with two fifths of children with ARFID exhibiting delayed language development at age 2.5—a well-established early indicator of neurodevelopmental conditions. Such difficulties may serve as early markers of elevated ARFID risk and could be stronger predictors than more typical early feeding deviations, although this warrants further investigation. Our findings suggest that the presence of ARFID-like feeding difficulties should prompt consideration of underlying neurodevelopmental concerns— and vice versa.

Furthermore, our study supports the utility of the ARFID-Brief Screener as an effective tool for identifying children at risk of ARFID. This parent-reported measure is easy to administer and may facilitate early identification in routine health check-ups, particularly among children with feeding challenges or neurodevelopmental difficulties. However, given the moderate specificity and positive predictive value, follow-up assessments remain necessary to confirm diagnoses and minimize false positives. Further studies are needed to validate our findings in other samples of preschool children.

## Data Availability

The datasets used and/or analysed during the current study are available from the corresponding author on reasonable request.

## Declarations

### Ethics approval and consent to participate

The study was approved by Swedish Ethical Review Authority (no. 2020-01284, 2020-03908, 2021-01849). All participants gave written informed consent. All methods were conducted in compliance with the applicable guidelines and regulations.

### Consent for publication

Not applicable

### Authors’ contributions

Conceptualization: Dinkler, Brimo, Holmäng, Yasumitsu-Lovell, Möllborg, Gillberg, Råstam Data curation: Dinkler, Brimo, Holmäng

Formal analysis: Dinkler

Funding acquisition: Dinkler, Yasumitsu-Lovell, Fujieda, Suganuma, Gillberg, Råstam Investigation: Dinkler, Brimo, Holmäng, Kantzer, Omanovic

Methodology: Dinkler, Kuja-Halkola, Bryant-Waugh, Råstam Project administration: Dinkler, Råstam

Resources: Dinkler, Brimo, Holmäng, Fernell, Möllborg, Råstam Software: Dinkler

Supervision: Bryant-Waugh, Gillberg, Råstam Validation: Eitoku

Writing–original draft: Dinkler

Writing–review and editing: All authors Visualization: Dinkler

Approved submitted version: All authors

## Acknowledgements

We thank the participants and the staff at the 21 involved child health services centers in West Sweden for their assistance with data collection.

## List of abbreviations

ARFID: Avoidant restrictive food intake disorder
AUC: Area under the curve
BMI: Body mass index
CHS: Child health services
CI: Confidence interval
ESSENCE-Q: Early Symptomatic Syndromes Eliciting Neurodevelopmental Clinical Examinations Questionnaire
ICC: Intraclass correlation coefficient
NPV: Negative predictive value
PARDI: Pica, ARFID, and Rumination Disorder Interview
PPV: Positive predictive value

