## Supplement for "Avoidant restrictive food intake disorder (ARFID) in Swedish preschool children: a screening study"

**Table S1.** ARFID–Brief Screener v2: Items, response options, and matching to DSM-5 criteria

| # | DSM-5 Criterion | Item | Response options |
|---|---|---|---|
| 1 | A0 - Avoidance or restriction of food intake | Do you think your child has or has had problems with eating characterized by avoidance or restriction of foods (i.e., that your child eats only a small range of foods or very little overall)? | No, never<br>Yes, now<br>Yes, earlier<br>*If yes:*<br>Started (age): ___<br>Stopped (age): ___ |
| 2 | | Has any health professional (at preschool/school, child health services, or other health care) said that your child has problems with avoidant or restrictive eating? | |
| Skipping rule: Do not continue if you replied "No, never" to both question 1 and 2. | | | |
| 3 | A1 - Significant weight loss (or failure to grow/gain weight) | Have your child's eating habits led to your child losing weight, not gaining weight, or not growing taller as they should? | No, never<br>Yes, now<br>Yes, earlier<br>*If yes:*<br>Started (age): ___<br>Stopped (age): ___ |
| 4 | A2 - Significant nutritional deficiency | Has any health professional said that your child has nutritional deficiencies due to their eating habits (e.g., vitamin or iron deficiency)? | |
| 5 | A3 - Dependence on enteral feeding or oral nutritional supplements | Has your child been prescribed dietary supplements containing vitamins and/or minerals to address nutritional deficiencies? | |
| 6 | | Has your child required high-calorie supplements (e.g., nutritional drinks) to be able to maintain or gain weight? | |
| 7 | | After the age of 6 months, has your child required tube feeding (food or fluid via a tube in the nose or into the stomach) to maintain proper nutritional status? | |
| 8 | A4 - Marked interference with psychosocial functioning | Do your child's eating habits negatively affect their functioning almost daily (e.g., in preschool/school, activities with family/friends)? | |
| 9 | Sensory aversion profile | Does your child often avoid eating foods with certain smell, taste, appearance, temperature, or consistency/texture (e.g., crispy or soft)? | |
| 10 | Concern about aversive consequences profile | Does your child often avoid eating foods because they are worried about e.g., choking, vomiting/being sick, tummy aches, diarrhea, or an allergic reaction? | |
| 11 | Lack of interest profile | Does your child often eat too little because of low interest in eating and/or low appetite? | |
| --- | C - Eating disturbance not attributable to weight/shape concerns | ***---- Not applied due to young age of the children in this study ----***<br>*Only if your child is 6 years or older:* Has your child ever restricted their eating because they wanted to lose weight or because they were afraid of gaining weight? |  |
| 12 | D – Eating disturbance not attributable to concurrent medical condition | ***---- Assessed but excluded from diagnostic algorithm[1] ----***<br>Do you suspect or know that your child's eating problems are primarily due to a medical condition or a mental disorder? | No<br>Yes<br>*If yes*, which one? |

DSM-5 Criterion B (eating disturbance not due to lack of available food or a culturally sanctioned practice) is not included in the ARFID–Brief Screener.

[1] Of children meeting the DSM-5 Criterion A (i.e., A0 + at least one of A1–A4), n=3 parents indicated that the child's eating problems were primarily due to a medical condition or a mental disorder. The indicated conditions were constipation, autism, and prematurity; however, none of these conditions exclude a diagnosis of ARFID.

**Table S2**: Sample characteristics by screening group.

| Characteristic | Overall N = 645 | (1) Screen-positive for current ARFID N = 42 | (2) Screen-positive for previous ARFID N = 25 | (3) Screen-negative with current eating problems N = 111 | (4) Screen-negative with previous eating problems N = 25 | (5) Screen-negative with no history of eating problems N = 442 | p-value[1] |
|---|---|---|---|---|---|---|---|
| **Sex** | | | | | | | 0.999 |
| Female | 319 (49.5%) | 21 (50.0%) | 13 (52.0%) | 55 (49.5%) | 12 (48.0%) | 218 (49.3%) | |
| Male | 326 (50.5%) | 21 (50.0%) | 12 (48.0%) | 56 (50.5%) | 13 (52.0%) | 224 (50.7%) | |
| **Age at screen (months)** | 38.0 (9.0), 20.4-59.1 | 39.2 (9.6), 24.4-51.4 | 36.9 (8.6), 28.4-49.6 | 38.0 (9.0), 28.4-51.3 | 34.8 (8.0), 29.3-49.6 | 38.2 (9.1), 20.4-59.1 | 0.828 |
| **Routine check-up** | | | | | | | 0.288 |
| 2.5 years | 371 (57.5%) | 21 (50.0%) | 16 (64.0%) | 64 (57.7%) | 19 (76.0%) | 251 (56.8%) | |
| 4 years | 274 (42.5%) | 21 (50.0%) | 9 (36.0%) | 47 (42.3%) | 6 (24.0%) | 191 (43.2%) | |
| **Current BMI** | 16.4 (1.5), 12.7-24.4 | 15.3 (1.4), 12.7-18.8 | 15.8 (1.0), 13.9-17.8 | 16.3 (1.3), 13.6-20.0 | 16.9 (2.0), 14.2-24.4 | 16.5 (1.4), 13.4-22.1 | <0.001 |
| **Respondent** | | | | | | | 0.941 |
| Mother | 538 (83.8%) | 36 (87.8%) | 22 (88.0%) | 92 (82.9%) | 22 (88.0%) | 366 (83.2%) | |
| Father | 93 (14.5%) | 4 (9.8%) | 3 (12.0%) | 16 (14.4%) | 3 (12.0%) | 67 (15.2%) | |
| Mother & Father | 11 (1.7%) | 1 (2.4%) | 0 (0.0%) | 3 (2.7%) | 0 (0.0%) | 7 (1.6%) | |
| **Mother born in Sweden** | 521 (80.8%) | 27 (64.3%) | 19 (76.0%) | 98 (88.3%) | 18 (72.0%) | 359 (81.2%) | 0.011 |
| **Father born in Sweden** | 511 (79.8%) | 27 (64.3%) | 17 (70.8%) | 94 (84.7%) | 13 (52.0%) | 360 (82.2%) | <0.001 |
| **Mother's education**[2] | | | | | | | 0.305 |
| Less than compulsory school (<9y) | 11 (1.7%) | 1 (2.4%) | 1 (4.0%) | 3 (2.7%) | 0 (0.0%) | 6 (1.4%) | |
| Compulsory school (9y) | 32 (5.0%) | 3 (7.1%) | 1 (4.0%) | 6 (5.5%) | 2 (8.0%) | 20 (4.6%) | |
| High school (12y) | 184 (28.7%) | 16 (38.1%) | 7 (28.0%) | 39 (35.5%) | 11 (44.0%) | 111 (25.3%) | |
| Further education (13-14y) | 145 (22.6%) | 9 (21.4%) | 8 (32.0%) | 24 (21.8%) | 2 (8.0%) | 102 (23.2%) | |
| University/college degree (15-17y) | 269 (42.0%) | 13 (31.0%) | 8 (32.0%) | 38 (34.5%) | 10 (40.0%) | 200 (45.6%) | |
| **Father's education**[2] | | | | | | | 0.094 |
| Less than compulsory school (<9y) | 14 (2.2%) | 2 (4.8%) | 0 (0.0%) | 1 (1.0%) | 0 (0.0%) | 11 (2.5%) | |
| Compulsory school (9y) | 45 (7.1%) | 5 (11.9%) | 3 (12.5%) | 6 (5.7%) | 2 (8.0%) | 29 (6.7%) | |
| High school (12y) | 286 (45.4%) | 14 (33.3%) | 14 (58.3%) | 64 (61.0%) | 12 (48.0%) | 182 (41.9%) | |
| Further education (13-14y) | 111 (17.6%) | 10 (23.8%) | 1 (4.2%) | 12 (11.4%) | 4 (16.0%) | 84 (19.4%) | |
| University/college degree (15-17y) | 174 (27.6%) | 11 (26.2%) | 6 (25.0%) | 22 (21.0%) | 7 (28.0%) | 128 (29.5%) | |
| **A0 criterion (parent)** | | | | | | | <0.001 |
| Yes, currently | 158 (24.5%) | 41 (100.0%) | 10 (40.0%) | 107 (96.4%) | 0 (0.0%) | 0 (0.0%) | |
| Yes, earlier | 38 (5.9%) | 0 (0.0%) | 13 (52.0%) | 2 (1.8%) | 23 (92.0%) | 0 (0.0%) | |
| No, never | 448 (69.6%) | 0 (0.0%) | 2 (8.0%) | 2 (1.8%) | 2 (8.0%) | 442 (100.0%) | |
| **A0 criterion (health professional)** | | | | | | | <0.001 |
| Yes, currently | 87 (13.6%) | 31 (75.6%) | 4 (16.0%) | 52 (48.1%) | 0 (0.0%) | 0 (0.0%) | |
| Yes, earlier | 23 (3.6%) | 0 (0.0%) | 11 (44.0%) | 5 (4.6%) | 7 (28.0%) | 0 (0.0%) | |
| No, never | 531 (82.8%) | 10 (24.4%) | 10 (40.0%) | 51 (47.2%) | 18 (72.0%) | 442 (100.0%) | |
| **Sensory avoidance**[3] | | | | | | | <0.001 |
| Yes, currently | | 26 (61.9%) | 8 (34.8%) | 52 (48.1%) | 2 (8.0%) | | |
| Yes, earlier | | 1 (2.4%) | 4 (17.4%) | 2 (1.9%) | 3 (12.0%) | | |
| No, never | | 15 (35.7%) | 11 (47.8%) | 54 (50.0%) | 20 (80.0%) | | |

| Characteristic | Overall N = 645 | (1) Screen-positive for current ARFID N = 42 | (2) Screen-positive for previous ARFID N = 25 | (3) Screen-negative with current eating problems N = 111 | (4) Screen-negative with previous eating problems N = 25 | (5) Screen-negative with no history of eating problems N = 442 | p-value[1] |
|---|---|---|---|---|---|---|---|
| **Lack of interest**[3] | | | | | | | <0.001 |
| Yes, currently | | 37 (88.1%) | 11 (45.8%) | 63 (58.3%) | 1 (4.0%) | | |
| Yes, earlier | | 2 (4.8%) | 6 (25.0%) | 2 (1.9%) | 5 (20.0%) | | |
| No, never | | 3 (7.1%) | 7 (29.2%) | 43 (39.8%) | 19 (76.0%) | | |
| **Concern about aversive consequences**[3] | | | | | | | <0.001 |
| Yes, currently | | 4 (10.0%) | 0 (0.0%) | 4 (3.6%) | 0 (0.0%) | | |
| Yes, earlier | | 0 (0.0%) | 2 (8.7%) | 3 (2.7%) | 2 (8.0%) | | |
| No, never | | 36 (90.0%) | 21 (91.3%) | 103 (93.6%) | 23 (92.0%) | | |

[1] Categorical variables: n (%); Pearson's Chi-squared test. Continuous variables: M (SD), Min-Max; Wilcoxon rank sum test. [2] "y" indicates total number of total school years. [3] Parents of screen-negative children with no history of eating problems (Group 5) did not respond to this question due to the skipping rule of the ARFID-Brief Screener.

**Table S3**. Neurodevelopmental problems measured by ESSENCE-Q separately by symptom in children with versus without ARFID diagnosis (dx) in the PARDI

| Characteristic | No ARFID dx in PARDI N = 39 | ARFID dx in PARDI N = 23[1] | p-value[2] |
|---|---|---|---|
| General development | | | 0.862 |
| No | 34 (87.2%) | 18 (81.8%) | |
| Maybe/a little | 2 (5.1%) | 2 (9.1%) | |
| Yes | 3 (7.7%) | 2 (9.1%) | |
| Motor development/milestones | | | >0.999 |
| No | 38 (97.4%) | 21 (95.5%) | |
| Maybe/a little | 0 (0.0%) | 0 (0.0%) | |
| Yes | 1 (2.6%) | 1 (4.5%) | |
| Sensory reactions (e.g. touch, sound, light, smell, taste, heat, cold, pain) | | | 0.039 |
| No | 37 (94.9%) | 16 (72.7%) | |
| Maybe/a little | 1 (2.6%) | 3 (13.6%) | |
| Yes | 1 (2.6%) | 3 (13.6%) | |
| Communication/language/babble | | | 0.543 |
| No | 31 (79.5%) | 16 (72.7%) | |
| Maybe/a little | 5 (12.8%) | 2 (9.1%) | |
| Yes | 3 (7.7%) | 4 (18.2%) | |
| Activity (overactivity/passivity) or impulsivity | | | 0.150 |
| No | 31 (79.5%) | 14 (63.6%) | |
| Maybe/a little | 6 (15.4%) | 3 (13.6%) | |
| Yes | 2 (5.1%) | 5 (22.7%) | |
| Attention/concentration/"listening" | | | 0.254 |
| No | 30 (76.9%) | 14 (60.9%) | |
| Maybe/a little | 7 (17.9%) | 5 (21.7%) | |
| Yes | 2 (5.1%) | 4 (17.4%) | |
| Social interaction/interest in other children | | | 0.315 |
| No | 34 (87.2%) | 17 (77.3%) | |
| Maybe/a little | 4 (10.3%) | 2 (9.1%) | |
| Yes | 1 (2.6%) | 3 (13.6%) | |
| Behavior (e.g. repetitive, routine insistence) | | | 0.245 |
| No | 32 (82.1%) | 15 (68.2%) | |
| Maybe/a little | 6 (15.4%) | 4 (18.2%) | |
| Yes | 1 (2.6%) | 3 (13.6%) | |
| Mood (depressed, elated/manic, extreme irritability, crying spells) | | | 0.084 |
| No | 31 (79.5%) | 12 (54.5%) | |
| Maybe/a little | 6 (15.4%) | 8 (36.4%) | |
| Yes | 2 (5.1%) | 2 (9.1%) | |
| Sleep | | | 0.666 |
| No | 32 (82.1%) | 17 (77.3%) | |
| Maybe/a little | 6 (15.4%) | 3 (13.6%) | |
| Yes | 1 (2.6%) | 2 (9.1%) | |
| Feeding | | | <0.001 |
| No | 2 (5.1%) | 0 (0.0%) | |
| Maybe/a little | 21 (53.8%) | 2 (8.7%) | |
| Yes | 16 (41.0%) | 21 (91.3%) | |
| "Funny spells"/absences | | | >0.999 |
| No | 36 (92.3%) | 21 (95.5%) | |
| Maybe/a little | 2 (5.1%) | 1 (4.5%) | |
| Yes | 1 (2.6%) | 0 (0.0%) | |

[1] One child had missing values on all ESSENCE--Q variables except on *Attention/concentration/"listening"* (Maybe/a little) and *Feeding* (Yes). [2] All variables: n (%), Pearson's Chi-squared test (all expected cell counts ≥5) or Fisher's exact test (any expected cell count <5).

**Table S4**. Agreement between ARFID-Brief Screener (Screen +/-) and clinical interview (PARDI +/-)

*Classification tables*

| Test No. | Ages | Definition of Screen- | | PARDI + | PARDI - |
|---|---|---|---|---|---|
| 1 | Across ages | Group 2 only | **Screen +** | 21 | 8 |
| | | | **Screen -** | 1 | 13 |
| 2 | Across ages | Group 3 only | **Screen +** | 21 | 8 |
| | | | **Screen -** | 1 | 18 |
| 3 | 2.5-year-olds only | Groups 2 & 3 | **Screen +** | 11 | 4 |
| | | | **Screen -** | 1 | 20 |
| 4 | 4-year-olds only | Groups 2 & 3 | **Screen +** | 10 | 4 |
| | | | **Screen -** | 1 | 11 |

*Statistical tests*

| **Test No.** | **1** | **2** | **3** | **4** |
|---|---|---|---|---|
| **Statistic** | **Estimate (95%CI)** | **Estimate (95%CI)** | **Estimate (95%CI)** | **Estimate (95%CI)** |
| Sensitivity | 0.95 (0.77-1.00) | 0.95 (0.77-1.00) | 0.92 (0.62-1.00) | 0.91 (0.59-1.00) |
| Specificity | 0.62 (0.38-0.82) | 0.69 (0.48-0.86) | 0.83 (0.63-0.95) | 0.73 (0.45-0.92) |
| Positive predictive value | 0.72 (0.53-0.87) | 0.72 (0.53-0.87) | 0.73 (0.45-0.92) | 0.71 (0.42-0.92) |
| Negative predictive value | 0.93 (0.66-1.00) | 0.95 (0.74-1.00) | 0.95 (0.76-1.00) | 0.92 (0.62-1.00) |
| Accuracy | 0.79 (0.64-0.90) | 0.81 (0.67-0.91) | 0.86 (0.71-0.95) | 0.81 (0.61-0.93) |
| F1 Score[1] | 0.82 (0.55-0.88) | 0.82 (0.65-0.91) | 0.81 (0.78-0.98) | 0.80 (0.62-0.95) |

[1] Bootstrapped confidence intervals
Screen +: screen-positive for current ARFID (group 1)
Screen -: screen-negative for current ARFID but with potential previous ARFID (group 2) or current eating problems (group 3)
PARDI +: ARFID diagnosis in PARDI
PARDI - : no ARFID diagnosis in PARDI

ARFID: avoidant restrictive food intake disorder; PARDI: Pica, ARFID, and Rumination Disorder Interview.
